# Threshold-based Identification of Persistent Infarction after Successful Endovascular Treatment: Re-evaluating the “Core”

**DOI:** 10.1101/2020.04.09.20048744

**Authors:** Raphael Meier, Paula Lux, Richard McKinley, Simon Jung, Urs Fischer, Jan Gralla, Roland Wiest, Johannes Kaesmacher

**Affiliations:** University Institute of Diagnostic and Interventional Neuroradiology, University Hospital Bern, Inselspital, University of Bern, Bern, Switzerland; Support Center for Advanced Neuroimaging, University Hospital Bern, Inselspital, University of Bern, Bern, Switzerland; Department of Neurology, University Hospital Bern, Inselspital, University of Bern, Bern, Switzerland; Department of Diagnostic, Paediatric and Interventional Radiology, University Hospital Bern, Inselspital, University of Bern, Bern, Switzerland

**Keywords:** Acute ischemic stroke, DWI, persistent infarction, reversal, ADC, threshold, endovascular treatment

## Abstract

**Objectives:** The objectives of this study included the volumetric analysis of persistent infarction and lesion reversal in Diffusion-Weighted Imaging (DWI), as well as the assessment of accuracy of ADC thresholds to identify regions of persistent infarction in patients with acute ischemic stroke after successful endovascular treatment (EVT).

**Materials and Methods:** A retrospective analysis of patients with M1 or proximal M2 occlusions, treated between 01/2012 and 07/2017, who underwent successful EVT (≥TICI 2b) and both pre- and post-interventional Magnetic Resonance Imaging (MRI), led to the inclusion of N=90 patients. Administration of recombinant tissue plasminogen activator (rTPA) for intra-venous thrombolysis was performed ahead of intervention in 45 cases (N=45/90, 50%). The majority of patients (N=78/90, 86.7%) were treated with second-generation thrombectomy devices with or without intra-arterial urokinase. DWI at admission and 24-hour follow-up DWI data were co-registered. Acute ischemic changes at baseline DWI, 24-hour DWI lesion, and the affected gray/white matter regions were manually annotated. Persistent infarction was defined as acute ischemic changes on baseline DWI, which were sustained on 24-hour follow-up DWI. Based on the manual annotations, persistent infarction and DWI reversal were quantified in a voxel-wise analysis. Thresholds for the identification of persistent infarction using baseline ADC images were estimated by maximizing Youden’s J statistic (ROC-analysis).

**Results:** Median age of the patients was 71.9 years (IQR 60.4–79.7 years), 55.6% were female, and NIHSS at admission was 11 (IQR 6–14). The median DWI lesion volume at baseline was 9.9 mL (IQR 3–23.6 mL) and the median DWI lesion volume around 24 hours was 12.1 mL (IQR 3.6–23.7 mL). Reversal of acute ischemic changes occurred frequently (49.8%, IQR 31.7%–65.4%; percentage of initial DWI lesion volume per subject). Sizeable DWI reversal (i.e. >10 mL and >10%) was observed in 26.7% (N=24/90) of the cases. Relative DWI reversal was significantly higher in white matter (58.6%, IQR 35.3–81.5%) than in gray matter (39.2%, IQR 24.9–56.6%; p<0.001). The volume of persistent infarction and DWI reversal were both significantly correlated with the DWI lesion volume at baseline (R=0.873–0.945, p<0.001), however, no correlations with time to reperfusion were found (relative volumes: R=-/+0.058, p=0.607). ROC analyses of ADC thresholds yielded optimal values which differed significantly for gray and white matter (p=0.003), and were lower than previously reported thresholds while having significantly improved accuracy (p≤0.015). No correlations between the estimated ADC thresholds and different covariates were found (time from imaging to reperfusion, time from baseline to follow-up imaging, volume of acute ischemic changes).

**Conclusions:** DWI reversal occurs frequently in successfully reperfused patients treated with modern EVT. Identification of persistent infarction using ADC thresholds in baseline DWI remains challenging with notable differences for gray and white matter.

## Introduction

Current imaging concepts of treatment selection for acute ischemic stroke patients rely on identification and delineation of irreversible ischemic brain tissue on admission imaging^2^. In case of Magnetic Resonance Imaging (MRI), this is usually realized using threshold-based segmentation of Apparent Diffusion Coefficient (ADC) maps derived from diffusion-weighted imaging (DWI). Best thresholds for identifying persistent infarction (ischemic core), despite restored reperfusion have been reported at ≤620 (μm^2^)/s^23^, with some studies reporting differences for gray and white matter, respectively^3,5,22^. Currently, there is paucity of data, if threshold-based identification of regions with persistent infarction is comparably accurate in case of rapid reperfusion by endovascular treatment (EVT). A recent pilot study suggested that ADC evolution in ischemic stroke patients with early endovascular reperfusion is strikingly different from findings observed in patients treated with intravenous thrombolysis (IVT)^11^. Specifically, early ADC normalization and core reversal was observed in more than 2/3 of patients undergoing endovascular treatment, therefore challenging the ADC cut-off derived from IVT treated patient cohorts^11^.

Within the present study, we performed a volumetric analysis of infarct evolution, including persistent infarction and DWI reversal, and assessed the accuracy of ADC thresholds to identify regions with persistent infarction in patients with successful endovascular treatment for a homogenous cohort of patients presenting with an M1 or proximal M2 occlusion at a tertiary care center.

## Methods

### Patients

We retrospectively analyzed consecutive mothership patients treated between 01/2012 and 07/2017 (N=720) by endovascular therapy. Indications for endovascular treatment, standardized imaging work-up, and mode of clinical follow-up have been published before^15^. Out of 720 patients, 474 patients received pre-interventional MRI, including 331 patients with M1 or proximal M2 occlusion. Of these, 120 patients underwent follow-up by MRI around 24 hours after reperfusion (median 23.4 h, IQR 20.5–26.5 h). We excluded patients without an acute lesion on admission DWI (N=5), patients with coarse co-registration errors (N=5) and patients with unsuccessful reperfusion (N=20). Hence, the final study population consisted of N=90 patients. The registry was approved by the local ethics committee (Kantonale Ethikkommission für die Forschung Bern, Bern, Switzerland, amendment access number: 231/2014). The raw patient-level data that support the findings of this study are available from the corresponding author on reasonable request and after clearance by the local ethics committee.

### Baseline and follow-up MRI

DWI sequences were acquired using a Magnetom Avanto or Magnetom Verio (Siemens, Erlangen Germany) at baseline and follow-up with two b values of 0 and 1000 s/mm^2^. Scanner parameters for DWI were slice thickness 5 mm, repetition time 3000 ms, echo time 89 ms, number of averages 4, flip angle 90°, voxel spacing (dx, dy, dz): 1.2×1.2×6.5 mm^3^ or slice thickness: 4 mm, repetition time 3500 ms, echo time: 89 ms, number of averages: 4, flip angle: 90°, voxel spacing (dx, dy, dz): 1.8×1.8×5.2 mm^3^, for 1.5 T and 3 T, respectively.

### Segmentation and image registration

Manual segmentation of regions of interest were performed by a medical master student (P.L.) using the simultaneous display of b=1000 s/mm^2^ and ADC images in the 3D Slicer Software^8^ (Version 4.8.0, https://www.slicer.org/) and by strictly following a standardized top-to-bottom approach of consecutive slices. Subsequently, the manual segmentations were reviewed and corrected by a neuroradiologist of >3 years clinical experience (J.K.). Acute ischemic changes were defined as hyperintensity in b=1000 s/mm^2^ images with corresponding hypointensity in the ADC image. Manual segmentation was performed accordingly to capture the maximum visual extent of the DWI lesion. Gray matter was segmented manually using the b=0 s/mm^2^ and 1000 s/mm^2^ images in combination with the ADC image. Manual segmentation of the DWI lesion and gray matter was performed for both baseline imaging and 24-hours follow-up in the same manner. Large confluent regions of interest were segmented using the Level Tracing Effect Tool of the Editor module. Manual correction or segmentation of small ischemic changes were outlined using the “Paint Effect Tool” of the same software. Small hemorrhagic transformations, discernible on susceptibility-weighted imaging, have been incorporated into the 24-hours lesion mask.

Subsequently, the follow-up ADC image was co-registered to the baseline ADC image using an affine co-registration algorithm (ITK Affine transform with linear interpolation using Mattes Mutual Information as optimization metric^12^). The estimated affine transform was used to map the label map of the follow-up DWI lesion to the image space of the baseline image. The co-registered images were visually checked for co-registration errors. A total of five patients were excluded due to coarse registration errors caused by image artifacts. Finally, an ADC threshold of >1500 (μm^2^)/s was applied to the lesion masks in order to exclude any voxel containing cerebrospinal fluid (csf)^3^.

Based on the co-registered manual segmentations, we defined three regions of interest: acute ischemic changes, persistent infarction, and DWI reversal. Acute ischemic changes correspond to the manual segmentation of the DWI lesion visible in baseline imaging. Persistent infarction was defined as acute ischemic changes which were contained in the follow-up lesion defined on the 24-hours ADC image. DWI reversal was defined as acute ischemic changes which were not contained anymore in the follow-up lesion. In conjunction with the manually segmented gray matter mask, we analyzed the three regions of interest with respect to their involvement of gray matter and white matter (=parenchyma outside of gray matter mask).

### Endovascular treatment

Institutional guidelines of acute stroke patient care and eligibility criteria for endovascular treatment are provided online (via this [link](http://www.neurologie.insel.ch/fileadmin/neurologie/neurologie_users/Unser_Angebot/Dokumente/Stroke-Guidelines-07-19-english.pdf)). Intra-venous thrombolysis with recombinant tissue plasminogen activator (rTPA) was administered ahead of intervention in 45 cases (N=45/90, 50%). Intra-arterial thrombolysis with urokinase^14^ as stand-alone therapy was used in four cases (N=4/90, 4.4%). Intervention was waived in eight cases (N=8/90, 8.9%) due to preinterventional reperfusion on DSA ahead of endovascular treatment. All other cases (N=78/90, 86.7%) were treated with second-generation thrombectomy devices with or without intra-arterial urokinase (>90% Solitaire or Capture, size 4–6/20–40 mm or 3/15 mm, respectively; Medtronic, Ireland). Endovascular procedures were typically performed without intravenous heparin through a 20-cm-long 8 or 9F sheath under local or general anesthesia, depending on patient cooperation and comorbidities. If technically feasible, a balloon-guiding catheter and/or a large-bore distal catheter was used for concomitant aspiration. Final modified Thrombolysis in Cerebral Infarction (TICI) grade was evaluated by an independent research fellow. If spontaneous reperfusion between baseline imaging and start of angiography has occurred, the time point of initial angiography runs was used as a proxy for the time-point of reperfusion.

### Statistical analysis

Descriptive statistics are provided as median (Interquartile Range, IQR) or frequency counts. Comparison of frequency counts were conducted applying Fisher’s exact test. Comparison of continuous or discrete variables with ordinal scale was performed using Mann-Whitney-U-Test for unpaired data and Wilcoxon signed-rank test for paired data. Receiver operating characteristic (ROC) analysis was performed for estimating optimal ischemic thresholds in baseline ADC images to identify persistent infarction on a per-subject basis. Computation of binary classification statistics was restricted to the region manually annotated as acute ischemic changes. If a voxel was labeled by a given threshold (ADC ≤value) and it corresponded to a labeled voxel in the 24-hours follow-up lesion, it was regarded as a true positive. If it was labeled by applying the threshold but did not correspond to a voxel in the follow-up lesion, it was rated as a false positive. A false negative, was a voxel not labeled by applying the threshold but labeled as lesion voxel in the 24-hours image. Finally, if a voxel was not labeled as a lesion voxel by applying the threshold and it corresponded to a non-lesion voxel in the 24-hours image, it was rated as a true negative. ROC analysis was conducted by starting with an initial ADC value of 0 (μm^2^)/s and incrementally increasing it by a value of 10 (μm^2^)/s up to a value of 1500 (μm^2^)/s. The optimal ischemic threshold was subsequently determined by maximizing Youden’s J statistic, which is equals to sensitivity + specificity - 1. Furthermore, sensitivity, specificity, positive predictive value (PPV), negative predictive value (NPV), and accuracy of the optimal ischemic thresholds are reported.

## Results

### Baseline characteristics

Ninety patients diagnosed with acute ischemic stroke of the anterior circulation and successful reperfusion (defined as TICI 2b or larger) were included in this analysis. Median age of the patients was 71.9 years (IQR 60.4–79.7 years), 55.6% were female, and NIHSS at admission was 11 (IQR 6–14). The median DWI lesion volume at baseline was 9.9 mL (IQR 3–23.6 mL) and the median DWI lesion volume around 24 hours was 12.1 mL, (IQR 3.6–23.7 mL). Figure 1 shows the manual segmentation of the acute ischemic changes, the DWI lesion in the 24-hours follow-up image, and the gray matter for an exemplary patient. An overview on relevant baseline characteristics of the patient cohort is given in Table 1 (for more detailed overview, see Supplementary Table I). Information on time from stroke onset to reperfusion was available for n=81 patients (median 235 min, IQR 177–339 min).

**Table 1.** Baseline characteristics of all included patients.

|  | Final patient population (N=90) |
| --- | --- |
| Age | 71.9 (60.4-79.7) |
| Sex, female | 55.6% (50/90) |
| Admission NIHSS | 11 (6-14) |
| TOAST |  |
| - Large-artery atherosclerosis | 11.1% (10/90) |
| - cardioembolic | 45.6% (41/90) |
| - other etiology | 7.8% (7/90) |
| - unknown etiology | 35.6% (32/90) |
| Witnessed symptom onset to first imaging (min) | 107 (82-145, n=60) |
| Last-Seen to first imaging (min) | 325 (138-556, n=25) |
| Thrombus length (mm) | 9.7 (7.5-15.2) |
| Occlusion Site |  |
| - Proximal M1 | 35.6 (32/90) |
| - Distal M1 | 41.1% (37/90) |
| - M2 | 23.3% (21/90) |
| Tandem | 13.3% (12/90) |
| IVT | 50% (45/90) |
| TICI 3 reperfusion | 56.7% (51/90) |
| 24h NIHSS | 2 (1-5, n=81) |

**Figure 1.**
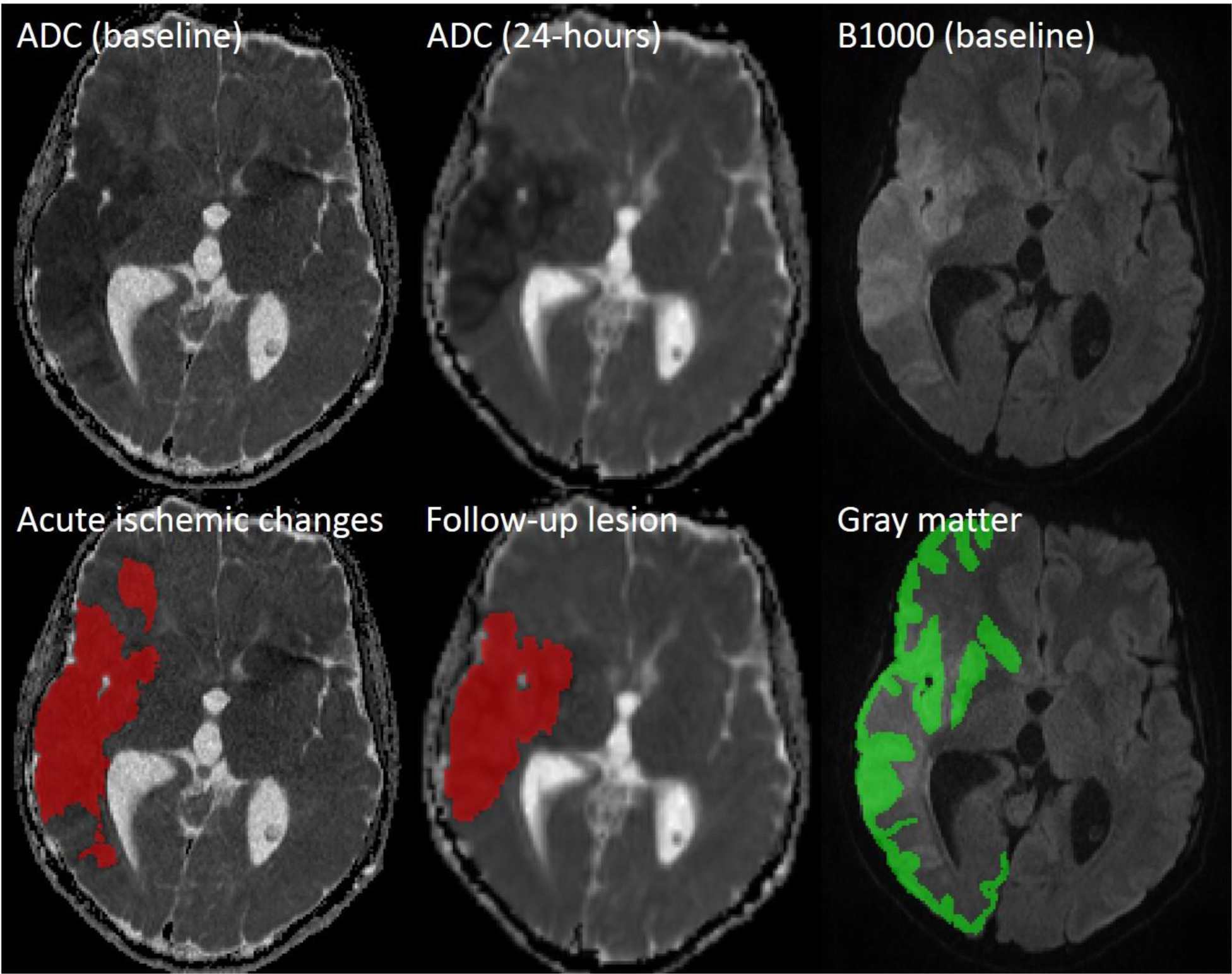
Exemplary patient case showing baseline ADC/B1000 image and co-registered 24-hours ADC. In the bottom row, manual segmentations of acute ischemic changes, follow-up lesion, and gray matter are overlaid.

To assess the direction and extent of bias, associated with confining our analysis to this subcohort, we compared baseline characteristics and outcomes of patients with and without 24-hours follow-up MRI (see Supplementary Table II). There was a general trend towards favorable baseline characteristics in patients receiving an early post-interventional MRI follow-up, as evidenced by younger age (75 vs 72 years, p=.029), lower admission NIHSS (10 vs 12, p<.001) and a more favorable risk factor profile.

### Infarct evolution and DWI reversibility

The volumetric measurements for acute ischemic changes, persistent infarction, and DWI reversal are shown in Figure 2. The median DWI lesion volume at baseline (9.9 mL, IQR 3– 23.6 mL) was significantly larger than the volume of persistent infarction (5.7 mL, IQR 0.9– 12.7 mL, p<0.001) around 24 hours. As shown in Figure 3, the volume of persistent infarction and DWI reversal were both significantly correlated with the DWI lesion volume at baseline (R=0.873–0.945, p<0.001). Furthermore, the volume of persistent infarction was significantly larger than DWI reversal for gray matter (3.5 mL, IQR 0.7–7.3 mL versus 1.4 mL, IQR 0.6–4.5 mL; p<0.001) and comparable for white matter (2.2 mL, IQR 0.4–5.1 mL versus 1.9 mL, IQR 0.7–5.9 mL; p=0.865).

**Figure 2.**
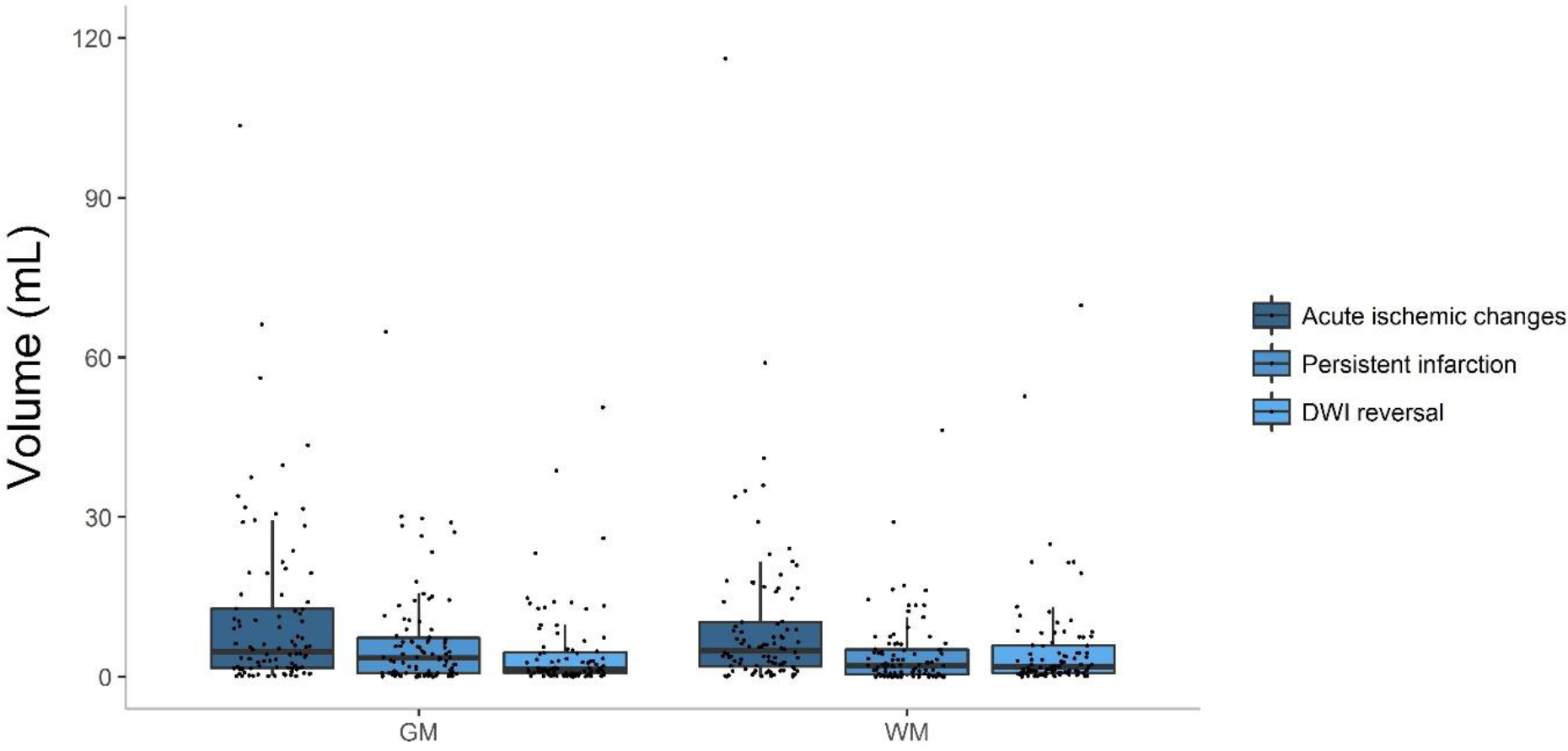
Volumetric measurements derived from manual segmentation including acute ischemic changes, persistent infarction, and DWI reversal.

**Figure 3.**
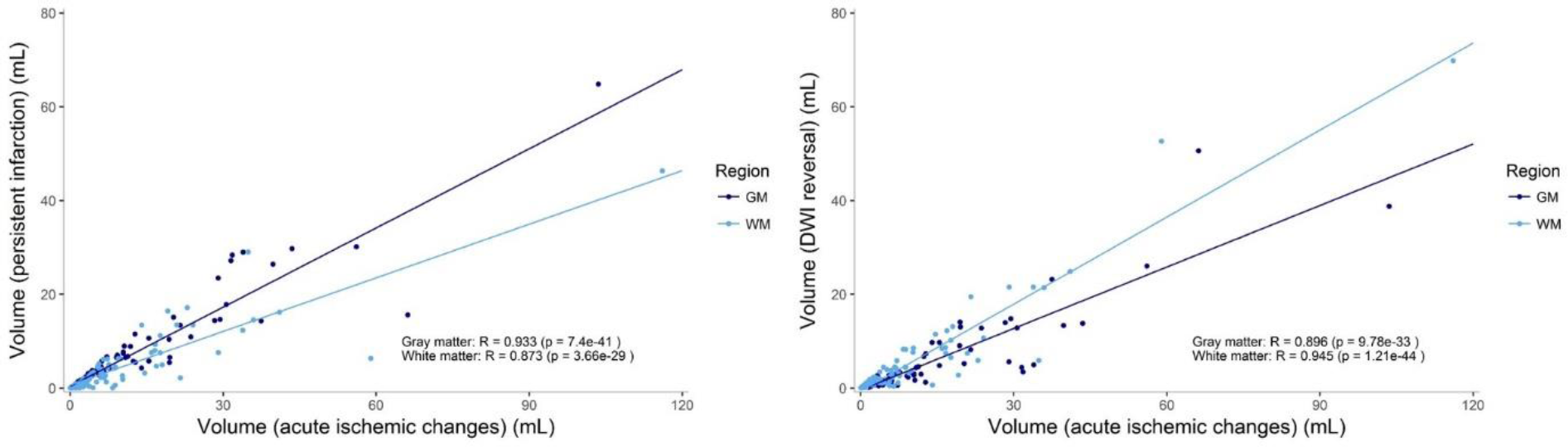
Scatter plot with linear correlation between volume of persistent infarction (left) or volume of DWI reversal (right) versus the volume of acute ischemic changes measured at baseline.

DWI reversal on a per-subject basis occurred frequently (49.8%, IQR 31.7%–65.4%; percentage of initial DWI lesion volume) with corresponding absolute volumes of 3.5 mL (IQR 1.4–10.9 mL). Sizeable DWI reversal (i.e. >10 mL and >10%) was observed in 26.7% (N=24/90) of the cases. When comparing DWI reversibility of gray and white matter, relative DWI reversal was significantly higher in white matter (58.6%, IQR 35.3–81.5%) than in gray matter (39.2%, IQR 24.9–56.6%; p<0.001) with comparable measurements for absolute volume (1.9 mL, IQR 0.7–5.9 mL versus 1.4 mL, IQR 0.6–4.5 mL; p=0.158).

When considering the relationship of relative persistent infarction and DWI reversal with time from stroke onset to reperfusion (TTRP, measured in minutes), no correlation was found (relative volumes: R=-/+0.058, p=0.607). Figure 4 shows relative and absolute volumes of persistent infarction and DWI reversal versus time from stroke onset to reperfusion.

**Figure 4.**
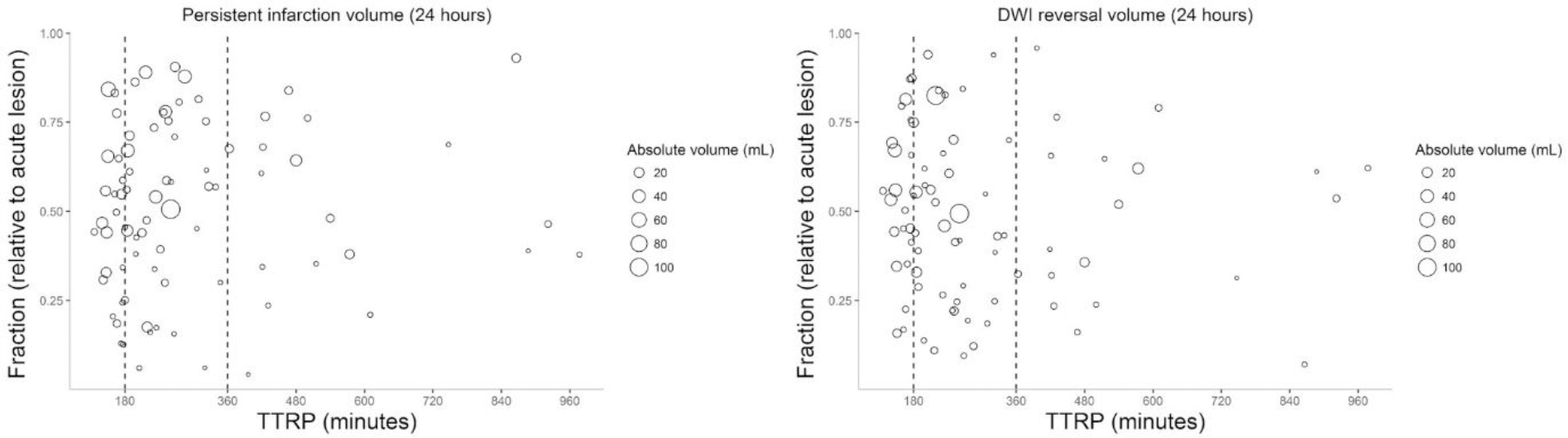
Scatter plot of relative persistent infarction (left) or relative DWI reversal (right) versus time from stroke onset to reperfusion (TTRP) in minutes. The size of the data point indicates the absolute volume of either persistent infarction or DWI reversal. The dotted vertical lines highlight the 3 hours and 6 hours marks.

The distributions of the subject-wise median ADC values for acute ischemic changes, persistent infarction, and DWI reversal differed significantly for both gray matter (p<0.001) and white matter (p<0.001) (see Figure 5). In particular, the subject-wise median ADC value for persistent infarction was significantly lower than the subject-wise median ADC value for DWI reversal for gray matter [508 (μm^2^)/s, IQR 479–542 (μm^2^) /s versus 564 (μm^2^)/s, IQR 524–595 (μm^2^)/s; p<0.001] and for white matter [494 (μm^2^)/s, IQR 457–529 (μm^2^) /s versus 549 (μm^2^)/s, IQR 515–577 (μm^2^)/s; p<0.001].

**Figure 5.**
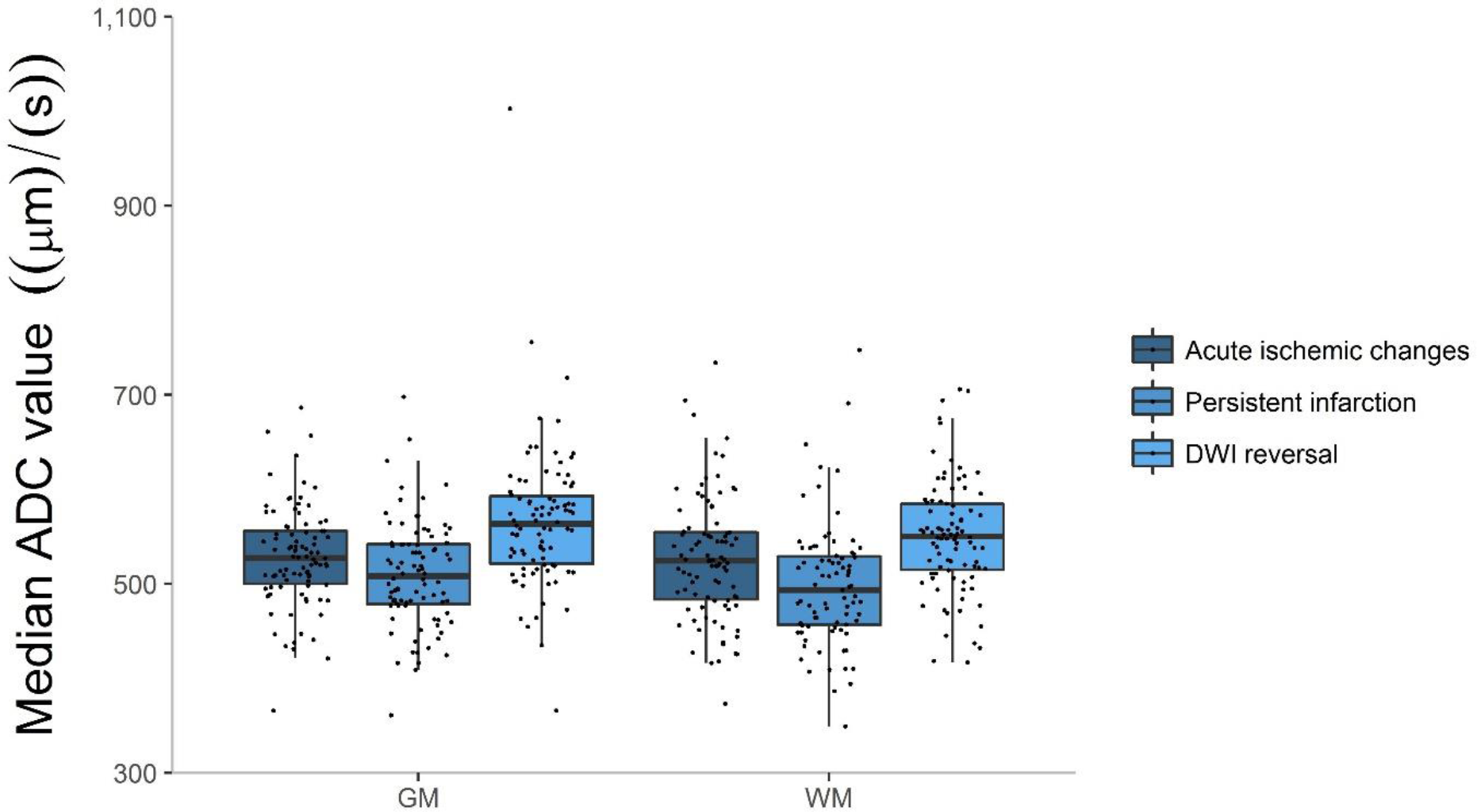
Boxplot of the subject-wise median ADC values for acute ischemic changes, persistent infarction, and DWI reversal.

When considering the relationship between relative persistent infarction and relative DWI reversal to the subject-wise median ADC, a weak negative linear association can be found in case of persistent infarction (gray matter: R=-0.219; p=0.038, white matter R=-0.421, p<0.001) and similarly a weak positive linear association can be found in case of DWI reversal (gray matter: R=0.219; p=0.038, white matter R=0.421, p<0.001) (see Figure 6).

**Figure 6.**
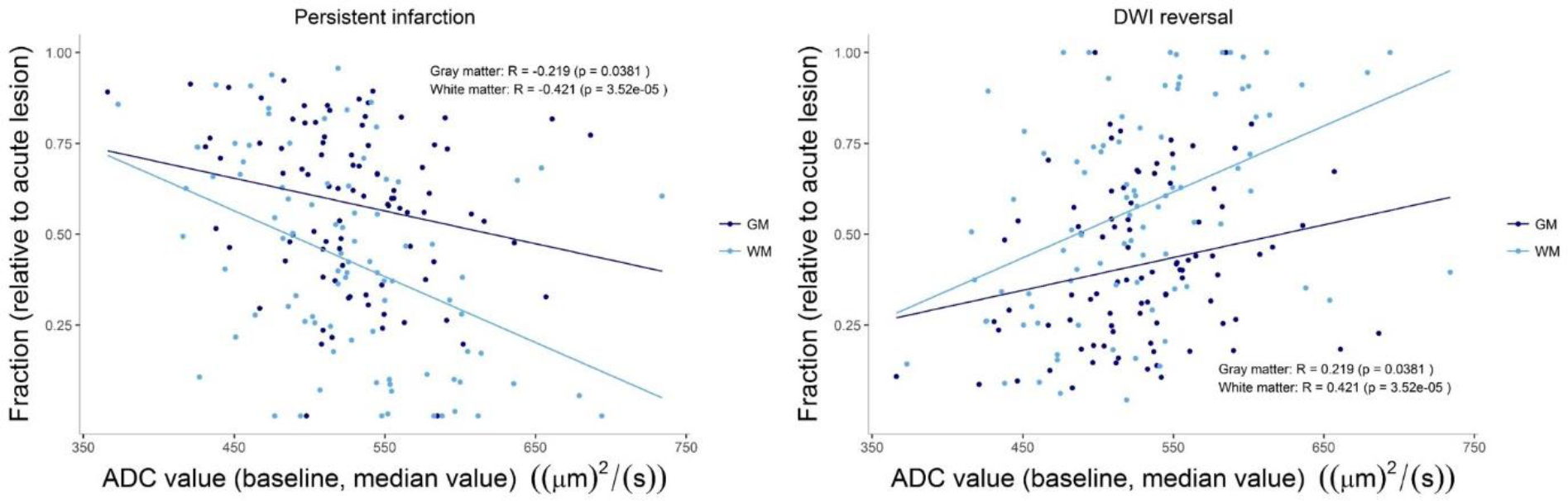
Scatter plot of relative persistent infarction (left) or relative DWI reversal (right) versus the subject-wise median ADC value.

### Ischemic threshold for persistent infarction

The application of the literature-based threshold of ADC ≤620 (μm^2^)/s for identifying the persistent infarction in a voxel-wise fashion resulted in a median accuracy value of 0.599 (IQR 0.505–0.698) for gray matter and a median accuracy value of 0.553 (IQR 0.401–0.652) for white matter. When performing subject-wise ROC analyses, the distribution of estimated ADC cutoff values yielded a lower median ADC cutoff value of 540 (μm^2^)/s (IQR 500–595 (μm^2^)/s) for gray matter and a lower median ADC cutoff value of 520 (μm^2^)/s (IQR 490–560 (μm^2^)/s) for white matter. In particular, the IQR of the two distributions of subject-wise ADC cutoff values lie consistently below the literature-based threshold of 620 (μm^2^)/s. Comparing the accuracy using the optimized subject-wise ADC cutoff values with the accuracy using the literature-based threshold, we observed a significant improvement for both gray matter (0.627 versus 0.599; p=0.015) and white matter (0.622 versus 0.553; p<0.001). For more details on the performance of ADC thresholds, see Table 2.

**Table 2.** Quantitative results of ischemic thresholds for identification of persistent infarction.

| Measurement:<br>Region: | Threshold ADC<br>($\mu\text{m}^2$)/s | AUC | Sensitivity | Specificity | PPV | NPV | Accuracy |
| --- | --- | --- | --- | --- | --- | --- | --- |
| Persistent infarction (GM) | 540 [500-595] | 0.623 [0.563-0.670] | 0.663 [0.559-0.760] | 0.623 [0.463-0.741] | 0.742 [0.569-0.844] | 0.545 [0.364-0.675] | 0.627 [0.582-0.696] |
| Persistent infarction (GM)<br>**fixed threshold** | **620** | - | 0.825 [0.714-0.887] | 0.321 [0.204-0.429] | 0.640 [0.464-0.788] | 0.490 [0.351-0.690] | 0.599 [0.505-0.698] |
| Persistent infarction (WM) | 520 [490-560] | 0.624 [0.582-0.705] | 0.655 [0.578-0.764] | 0.617 [0.510-0.750] | 0.575 [0.375-0.774] | 0.664 [0.469-0.874] | 0.622 [0.587-0.673] |
| Persistent infarction (WM)<br>**fixed threshold** | **620** | - | 0.900 [0.829-0.962] | 0.207 [0.135-0.332] | 0.483 [0.278-0.698] | 0.674 [0.482-0.897] | 0.553 [0.401-0.652] |

When performing pairwise comparison of subjects (n=79/90) who exhibited persistent infarction in both gray and white matter, consistently higher ADC cutoff values for gray matter than for white matter (p=0.003) were observed.

Subsequently, linear dependencies between time from imaging to reperfusion, time from baseline to follow-up imaging, volume of acute ischemic changes and the identified optimal ADC cutoffs and corresponding accuracy values were investigated. No significant correlations were found between the estimated ADC thresholds and different covariates (see Figure 7) as well as for the associated accuracy values and different covariates. There is a tendency for lesions exhibiting small acute ischemic volumes to have a larger variability in the estimated ADC thresholds and accuracy values (Figure 7, bottom row).

**Figure 7.**
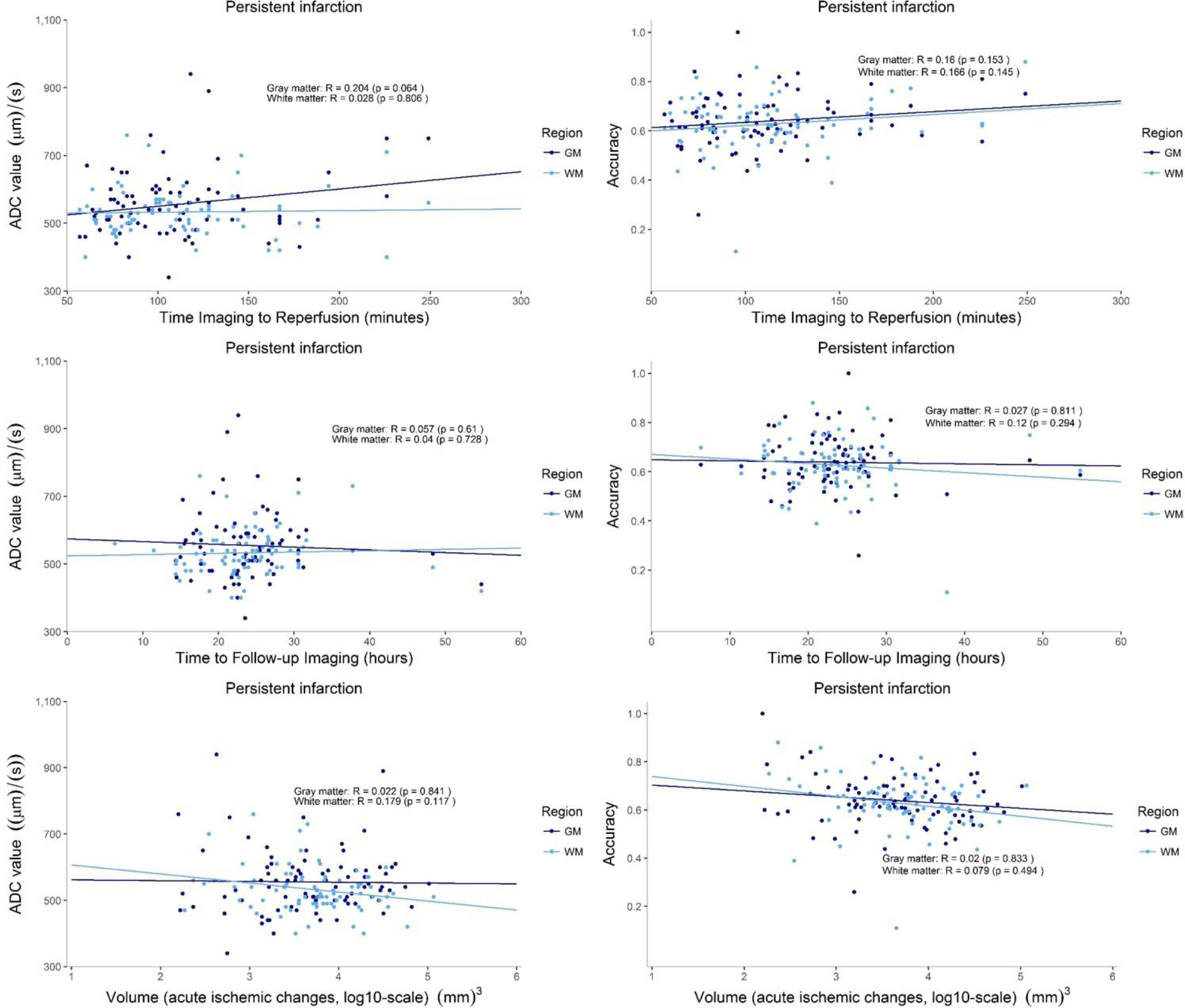
Scatter plots of optimal subject-wise ADC thresholds and corresponding accuracy values versus time baseline imaging to reperfusion (top row), time baseline imaging to follow-up imaging (middle), and volume of acute ischemic changes (bottom row).

## Discussion

Acute ischemic changes in patients undergoing successful EVT can either persist or reverse. When looking at the infarct evolution within 24 hours after EVT, we identified three major findings: (1) acute ischemic changes showed a tendency to persist in gray matter and to reverse in white matter (rel. DWI reversal of 39.2% versus 58.6%), (2) DWI reversal occurred frequently and was sizeable in about one quarter of the patients, (3) identification of persistent infarction required subject-wise optimal ADC thresholds which were below the literature-based threshold^23^ of 620 (μm^2^)/s with significantly improved accuracy and distinct differences for gray and white matter.

The appearance of ischemic tissue imaged at 24 hours after baseline imaging, can be described as distinctively dark on ADC and bright on DWI images. There are a number of previous studies which investigated DWI reversal in patients with acute ischemic stroke within 24 hours after baseline imaging^11,18,26,30^. A recent study by Hsia et al.^11^ found that the inflection point of ADC renormalization for the ischemic core after successful reperfusion may occur as early as 24 hours. Moreover, the issues of extensive tissue swelling and subsequent atrophy, which would further complicate image co-registration, can be avoided at 24 hours. Therefore, we focused on assessing infarct evolution and DWI reversibility using the 24-hours follow-up imaging. Furthermore, we can broadly differentiate between a subtraction-based analysis in which lesion volumes for two time points are computed and then subtracted to yield a change (e.g. lesion reversal), and a voxel-wise analysis which typically relies on a temporal co-registration of imaging data to yield a definition of voxel-wise change (e.g. lesion reversal). In contrast to a voxel-wise analysis, subtraction-based analysis has shown the tendency to misrepresent and mask the underlying DWI lesion dynamics in the past^10^. Therefore, we decided to conduct a voxel-wise analysis of DWI lesion dynamics using co-registered baseline and 24-hours follow-up imaging.

DWI reversibility of the ischemic core in stroke patients undergoing therapy with intravenous thrombolysis or mechanical thrombectomy has been investigated in the past. Going back more than 10 years, Kranz et al.^17^ performed a systematic review of literature, which at that time found DWI lesion reversal (defined as ≥10% decrease in lesion volume) in around 24% of the pooled patients. Based on this result, the authors hypothesized that in the first 24 hours after stroke onset, DWI hyperintensity represent a combination of both reversible and irreversible ischemia. A series of studies looking at DWI reversal in thrombolysis & early thrombectomy patients (EPITHET, DEFUSE 1/2 data) found that it was a rare phenomenon, confounded by CSF^7^, and being often transient (when evaluated on follow-up imaging < 12 hours)^6,13^. The results of these studies support the notion of an ischemic core. These results were complemented by other investigations, which found that while complete DWI reversal was rare, reversal of individual DWI hyperintensities was less rare^1^, and that reversal was significantly greater in patients treated within 3 hours^18^. Furthermore, some studies found DWI reversal (within 24 hours) to be associated with favorable clinical outcome^18,19^, while others did not^1^. More recent studies on thrombolysis patients observed that over two-thirds of the DWI lesion reversal captured on 24-hours DWI was sustained and reflected a strong correlate of early neurological improvement^26^, and that reversal frequently occurs in successfully recanalized patients with large cores (≥70 mL)^27^. Very recently, Yoo et al.^30^ found in a cohort of stroke patients treated with thrombectomy DWI reversal to occur in roughly 16% of patients. Moreover, timely reperfusion and complete reperfusion were associated with increased DWI reversal. The authors estimated DWI reversal in a subtraction-based fashion as decrease of volume from baseline to follow-up DWI. This may explain why in our study, which includes a voxel-wise analysis and only successfully reperfused patients, we observed a substantially larger frequency of DWI reversal. Hsia et al.^11^ investigated DWI reversal within 24 hours in a cohort of patients treated either with EVT +/- IVT or IVT alone. They defined significant reversal if > 50% of the baseline core was >620 (μm^2^)/s at 24 hours and found this to be the case for 69% of the successfully reperfused patients. Similar to our findings, this result indicates that sizeable DWI reversal is observed with increased frequency when looking at patients who achieved timely and successful reperfusion using modern EVT.

There have been several studies which aimed at characterizing ischemic thresholds in DWI of acute ischemic stroke patients. Looking back more than a decade, Meng et al.^21^ found an ischemic threshold of ADC ≤530 (μm^2^)/s in a rodent stroke model for middle cerebral artery occlusion. In human patients, ischemic thresholds for gray and white matter were found to be significantly different in two previous studies^3,5^. The studies differed in the way they defined acute ischemic changes with the first study by Bristow et al.^5^ defining it via abnormal regions in the mean-transit time perfusion maps and the second study by Arakawa et al.^3^ looking at hyperintense DWI with corresponding ADC ≤800 (μm^2^)/s. Furthermore, persistent infarction was defined once using 30-day follow-up imaging and once 90-day follow-up imaging. Both studies found significantly higher ADC thresholds for persistent infarction in gray matter than in white matter, which agrees with our results. However, the ROC analyses of both studies provided very different ischemic thresholds with Bristow et al. reporting ADC values of ≤786 and 708 (μm^2^)/s and Arakawa et al. reporting ADC values of ≤918 and 805 (μm^2^)/s for persistent infarction in gray and white matter, respectively. The study by Purushotham et al.^23^ determined the now widely-used ADC threshold of ≤620 (μm^2^)/s based on data of the DEFUSE study. They included 14 subjects with baseline DWI and 30-days follow-up imaging for which they determined the optimal ADC threshold of ≤620 (μm^2^)/s to discriminate persistent infarction from DWI reversal with an overall sensitivity of 69% and specificity of 78%. For determination of the optimal ADC threshold, they relied on the manual segmentation of the maximal visual extent of DWI abnormalities in baseline imaging of partially or completely recanalized patients, which corresponds to our methodology. Since their analysis was based on patients treated with thrombolysis, we hypothesize that the significantly lower optimal ADC thresholds observed in our analysis (ADC ≤520 and 540 (μm^2^)/s) might come from the faster and possibly more complete recanalization achieved through modern EVT techniques. This hypothesis is supported by data of a recent case control study which looked at the threshold for relative cerebral blood flow (rCBF) in CT Perfusion to define persistent infarction (ischemic core)^4^. They found that in patients with complete reperfusion treated by thrombectomy the optimal rCBF threshold is significantly lower than for thrombolysis patients with complete reperfusion—likely attributed to earlier reperfusion. Finally, we have to keep in mind that thresholds based on data from different patient cohorts under varying clinical and imaging conditions, will inevitably have limited generalizability. Sah et al.^25^ demonstrated in a subtraction-based analysis that lesion volumes and changes in lesion volumes over time for stroke patients differed significantly based on the chosen threshold for relative DWI intensity and absolute ADC. Therefore, based on the more recent evidence one might argue whether a threshold-based definition of persistent infarction in baseline DWI is a useful imaging criterion to start with. Recently, machine learning-based algorithms have been proposed for the segmentation of persistent infarction from MRI data, which may serve as an alternative to threshold-based algorithms^9,20,28^ in the near future. Furthermore, advances in MRI technology, such as e.g. diffusion kurtosis imaging, may potentially enable to discern differently injured portions in DWI lesions. In particular, it has been shown that kurtosis-based lesions are more likely to persist, whereas the kurtosis-diffusion mismatch can help in the identification of infarcted tissue which is likely to reverse after early reperfusion^29^.

The observed results have many clinical implications. First, current thresholds used in clinical routine will most likely overestimate the volume of persistent infarction when reperfusion is successful and occurs rapidly after the initial imaging. Second, current concepts of imaging selection do not distinguish between gray and white matter infarcts thereby preferably overestimating white matter infarcts. This may not only contribute to general overestimation of the core, but also may prompt individualized exclusion of patients in scenarios of white matter DWI hyperintensities in highly eloquent areas, such as lesions commonly observed in basilar artery occlusion. In addition, it has been shown that late white matter salvage is associated with good outcomes^16^, and according to our results, such a salvage may often be possible despite early DWI changes. Third, even considering the contemporary gold standard of infarct detection and segmentation, accuracy values for identifying areas of persistent and reversing ischemic changes seem suboptimal, and most certainly too low for planning individualized treatment decisions based on brain eloquence considerations. Lastly, there are ongoing efforts to identify areas of early ischemic changes on native CT. Recently published machine learning algorithms are usually trained on DWI ground-truth data acquired just before or after the native CT^24^. Although promising, the results here imply that these trained models are inherently associated with the same limitations and potential for misclassification as DWI itself.

The limitations of our study include the stringent inclusion of patients with baseline and follow-up MRI, which resulted in a homogenous cohort of patients with relatively small lesion volumes at baseline imaging. This selection bias of our cohort is potentially important, because patients with good clinical outcomes tend to have better collaterals, decreased infarct growth and more DWI reversal and hence, observed frequencies in this analysis may not be generalizable to other cohorts. Moreover, admission and follow-up imaging were not necessarily acquired at the same scanner or given the same magnetic field-strength, which leads to an overall reduced precision of image co-registration and resampling (shown in Supplementary Figure I for the follow-up lesion volume). Lastly, gray matter masks were manually segmented. While this clearly yields a segmentation which is maximally adjusted for the individual patient anatomy, it leaves uncertainty to inter-rater reliability and reproducibility.

In conclusion, we found that DWI reversal in successfully reperfused patients treated with modern EVT is a frequently occurring phenomenon, with roughly one quarter of the patients demonstrating sizeable DWI reversal. Characterization of persistent infarction required a markedly lower ADC threshold than has been proposed previously for thrombolysis patients with notable differences in infarct evolution for gray and white matter.

## Data Availability

The raw patient-level data that support the findings of this study are available from the corresponding author on reasonable request and after clearance by the local ethics committee.

